# Tongue swab testing on two automated tuberculosis diagnostic platforms, Cepheid Xpert® MTB/RIF Ultra and Molbio Truenat® MTB Ultima

**DOI:** 10.1101/2023.10.10.23296833

**Authors:** Rachel C. Wood, Angelique K. Luabeya, Rane B. Dragovich, Alaina M. Olson, Katherine A. Lochner, Kris M. Weigel, Renée Codsi, Humphrey Mulenga, Margaretha de Vos, Mikashmi Kohli, Adam Penn-Nicholson, Mark Hatherill, Gerard A. Cangelosi

## Abstract

Tongue dorsum swabbing is a potential alternative to sputum collection for tuberculosis (TB) testing. Previous studies showed that Cepheid Xpert® MTB/RIF Ultra (Xpert Ultra) can detect *Mycobacterium tuberculosis* (MTB) DNA in tongue swabs stored in buffer, with 72% sensitivity and 100% specificity relative to a sputum microbiological reference standard (sputum MRS). The present study evaluated a more convenient sample collection protocol (dry swab storage), combined with streamlined sample processing protocols, for side-by-side analysis using two commercial TB diagnostic tests: Xpert Ultra and Molbio Truenat® MTB Ultima (MTB Ultima). Copan FLOQSwabs were self-collected, or collected by study workers, from 321 participants in Western Cape, South Africa. All participants had symptoms suggestive of TB, and 245 of them had sputum MRS-confirmed TB (by sputum culture and/or Xpert Ultra). One tongue swab per participant was tested on Xpert Ultra and another tongue swab was tested with MTB Ultima. Xpert Ultra was 75.4% sensitive and 100% specific, and MTB Ultima was 71.6% sensitive and 96.9% specific, relative to sputum MRS. When sample lysates that were false-negative by MTB Ultima were frozen, thawed, and re-tested, MTB Ultima sensitivity rose to 79.1%. Both tests were more sensitive with swabs from participants with higher sputum Xpert semi-quantitative results. The protocol for Xpert Ultra enabled fast and easy testing of dry-stored swabs with no loss of accuracy relative to previous methods. MTB Ultima testing of dry-stored swabs exhibited comparable performance to Xpert Ultra. These results further support tongue swabs as easy-to-collect samples for high-throughput TB testing.

## INTRODUCTION

Tuberculosis disease (TB), caused by *Mycobacterium tuberculosis* (MTB), remains a major global cause of morbidity and mortality (1). The standard patient sample for microbiological diagnosis of TB is sputum, a viscous material expectorated from airways. The availability of alternative, noninvasive samples, which could be collected outside of a clinic, would greatly facilitate testing for active TB in community settings such as schools, workplaces, and institutions (2-4).

Researchers have sought sputum alternatives for years, but few materials have been identified that are both 1) easier than sputum to collect; and 2) reasonably accurate, relative to sputum testing (2, 3). To address this need, we and others have found that MTB cells and/or DNA can be detected by oral swab (OS) testing (5-17). In TB OS testing, the dorsum of the tongue is gently brushed with a swab for at least 10 seconds. The material collected by the swab is then subjected to nucleic acid amplification testing (NAAT) targeting MTB DNA. Tongue swabbing is fast, painless, and does not require specific infrastructure for privacy or infectious aerosol control. Everyone, including children and people living with HIV (PLWH), can be easily swabbed in any setting (18, 19).

Early evaluations of TB OS (9-11), as well as some more recent studies (5, 7, 8, 14), used in-house manual quantitative PCR (qPCR) methods to detect MTB DNA in tongue swab samples. Other evaluations used the commercial and WHO-approved Cepheid Xpert® MTB/RIF Ultra automated qPCR system (Xpert Ultra). Most evaluations of Xpert Ultra testing of tongue swabs used sample preparation protocols that mimicked those used for Xpert Ultra testing of sputum (12, 13, 17, 20). These approaches often yielded modestly accurate results when the sensitivity and specificity of OS were calculated relative to the sputum microbiological reference standard (sputum MRS). In a collaborative study conducted in Uganda, we evaluated new swab sample processing protocols that were designed explicitly for testing swabs in Xpert Ultra (6). These approaches yielded better accuracy, with 72% sensitivity relative to sputum MRS (incorporating both sputum Xpert Ultra and sputum culture), and 78% sensitivity relative to sputum Xpert Ultra alone. Specificity was 100% relative to these reference standards (6).

This study builds upon our previous work (6) in several ways. First, the previous study used swabs that were collected (either singly or in tandem) into 2-mL storage/transport tubes that were pre-filled with a sterile buffer, whereas this study used single swabs that were collected, cold-stored, and transported dry in 2-mL tubes, without buffer. Dry collection and storage may be more user-friendly for patients and providers, especially in community (non-clinical) settings (19). Second, the current study used a streamlined protocol for swab sample processing (about 15 minutes from sample tube to start of test, compared to 45 to 60 minutes for the methods described previously (6)). Third, the current study evaluated an alternative commercial TB diagnostic test, the Molbio Truenat® MTB Ultima (MTB Ultima) (3), applied to additional tongue swabs collected from the same participants. Finally, with 245 participants with confirmed TB by sputum MRS (culture and/or Xpert Ultra), this is the largest sensitivity study reported to date.

To our knowledge this is the first clinical evaluation of dry-stored tongue swabs for non-invasive, non-sputum TB testing, and the first side-by-side evaluation of Xpert Ultra and MTB Ultima applied to tongue swabs.

## METHODS

### Study setting

The study was conducted in Worcester, located in the Cape Winelands of the Western Cape province of South Africa. Tuberculosis (TB) burden in this area is high with a TB incidence rate of 700 per 100 000.

### Study Participants

Participants were recruited at the local TB clinics in two cohorts with distinct study designs. Cohort 1 (April 2021 – February 2022) was designed to study the impacts of patient oral hygiene behaviors and food /drink intake on OS testing results (21), and consisted solely of TB patients confirmed by sputum Xpert Ultra (N = 100). Sampling was split into 3 cases: Case 1 swabs were collected prior to eating/drinking or performing oral hygiene; only Case 1 samples were included in this analysis.

Cohort 2 (July 2021 – March 2023) was designed to assess OS analytical methods. All participants were asked not to eat or drink or perform any oral hygiene for at least 30 minutes prior to providing samples. Cohort 2 had two phases. In the first phase we enrolled patients with symptoms suggestive of TB, prior to confirmation of their disease. After enrollment of 96 participants, we observed that most (N = 79/96) did not have TB confirmed by MRS. Therefore, to meet target numbers of enrollees with confirmed TB, the Cohort 2 protocol was amended to enroll only people with sputum Xpert-confirmed TB.

This project was approved by the University of Cape Town Human Research Ethics Committee (reference number 160/2020) and the University of Washington Human Subjects Division (STUDY00001840).

### Sample collection

Swabs were Copan FLOQSwabs (520CS01), which are sterile, single-use, and individually packaged, with a break point at 30 mm. All swabs were collected within three days of initiation of TB treatment.

During their first visit, Cohort 1 participants provided oral swabs if they met the oral hygiene criteria of the first Case to which they were assigned. If they did not meet the criteria, their first oral swabs were collected during a subsequent visit to the participant’s home. During the participant’s Case 1 visit, 6 FLOQswabs were collected.

Cohort 2 participants were brought to the research site on Day 1 for informed consent, oral swabs, and sputum sample collection. On Day 1, five swabs were collected at the research site by the study team. On Day 2, two more swabs were self-collected by participants at their homes. The first two Day 1 swabs and the two Day 2 swabs were designated for assessment of OS analytical methods (including this study). Additional Day 1 swabs were saved for secondary analyses or for use as backups if instrument errors occurred.

In both Cohorts, oral swabs were collected by study workers if the swabbing took place at the clinic or on site at the South African Tuberculosis Vaccine Initiative (SATVI) offices.

Participants swabbed themselves under the supervision of study workers during home visits. The protocols for provider-collected swabs and supervised self-swabbing are provided elsewhere (19). After the swab was collected, either the study worker or the participant placed the swab into an empty 2-mL screw-cap tube (Sarstedt, 72.694.106) held by the study worker. The worker then broke or cut off the head of the swab into the tube and discarded the shaft. The tube was closed and placed into storage at -80 °C within 12 hours of collection. The samples were stored at -80 °C and shipped on dry ice. Upon receipt in Seattle, they were immediately transferred to the -80 °C freezer where they were stored until testing.

### Sample analysis

Prior to sample analysis, swab samples from both cohorts were randomized by collection day and collection order between the two test platforms, to prevent bias. Testing of Cohort 2 samples was blinded as to TB status.

The Molbio Truenat® MTB Ultima chip was run on the Molbio Truelab® machine, executing an automated PCR targeting IS*6110* and IS*1081* (3). Prior to the PCR assay, the swab samples had to be processed to render them safe to handle, and to release MTB DNA from bacilli. This was accomplished as follows. The wells of a heat block were filled with water and it was pre-heated to 100 °C. The water facilitated heat distribution. After pre-heating, the samples were removed from the freezer and immediately placed into the heat block where they were heated for 10 minutes. Samples were removed from the heat block and 500 µL TE buffer was added to the tubes, followed by vortexing (Vortex-Genie 2, SI-0236) for 15 seconds.

Samples were returned to the heat block and heated to 100 °C for another 10 minutes to facilitate lysis and elution of cells/DNA off the swab. Samples were then removed, briefly centrifuged, and allowed to cool for 5 minutes. Glass disruption beads (150 mg RPI 0.1 mm, pre-weighed and stored in individual tubes) were added, and samples were bead-beaten at max speed for 10 minutes on a vortexer using a vortex adapter (Mo Bio, 13000-V1-24). After a brief spin (5 seconds in a mini-centrifuge), the lysate was removed and transferred to a labeled 1.5-mL snap-cap tube. Six microliters of this lysate was run on the MTB Ultima chip, following the manufacturer’s instructions. Two Truelab® Duos were used to run the samples, with a capacity for running 2 samples each at a time. The lysis protocol for 4 samples took about 45 minutes, while the MTB Ultima assay took about 40 minutes.

This manual lysis protocol is considered off-label, as Molbio recommends using their Trueprep® AUTO v2 cartridge-based sample prep device for sample lysis and concentration. The Trueprep® cartridges contain an internal positive control (IPC) that is added to each sample during processing. Since we did not use the Trueprep®, there was no IPC in our samples. Instead, we monitored the performance of the MTB Ultima chips and Truelab® with a designated positive control from the Truenat® Positive Control Kit – Panel 1 after every 20 runs. If the positive control failed, the preceding 20 runs were considered not valid.

In the Xpert Ultra assay, cell lysis and PCR are conducted entirely within the Xpert Ultra cartridge. Therefore, the sample preparation protocol for Xpert could be shorter, taking an estimated 15 minutes per set of 4 samples. As in the Molbio protocol, the swabs were heated at 100 °C for 10 minutes in a water-filled heat block. Tubes were removed from the heat block and 1 mL TE buffer was added, followed by vortexing for 30 seconds. The 1-mL sample was transferred to the Xpert Ultra cartridge. Another 1 mL of TE was added to the original tube containing the swab. After another 30 second vortex, this additional 1-mL sample was added to the Xpert Ultra cartridge. The cartridge was then loaded into a 4-module GeneXpert instrument for analysis. If an error occurred during the Xpert test, a backup swab was selected, and the testing was repeated.

## RESULTS

### Study population characteristics

Three hundred and thirty participants (N = 330) were enrolled and N = 321 were included in the analysis. Cohort 1 had N = 100 participants. Cohort 2 had N = 222, of whom N = 145 were TB-positive patients by sputum MRS (sputum culture and/or Xpert Ultra positive), and N = 76 TB-negative patients. Some participants were excluded from the analysis due to incomplete sample collection. **Table 1** shows their characteristics, with Cohort 2 divided between TB positive and TB negative participants as defined by sputum MRS.

**Table 1.** Baseline characteristics of participants recruited for cohort1 and cohort 2.

| Variables | Cohort 1 (sputum TB positive) | Cohort 2 (sputum TB positive) | Cohort 2 (sputum TB negative) |
| --- | --- | --- | --- |
| N | 100 | 145 | 76 |
| Age (median, IQR) | 34 (27-44) | 35(25-45) | 38.5 (31.5-47.4) |
| Gender (Male) | 63(63%) | 86 (59.3%) | 31 (40.8%) |
| Mixed-race ancestry | 77 (77.0%) | 106 (73.1%) | 52 (68.4%) |
| Black | 23 (23.0%) | 38 (26.2%) | 22 (28.9%) |
| White | 0 | 1(0.7) %) | 1 (1.3%) |
| Asian | 0 | 0 | 1 (1.3%) |
| Employed | 31 (31.0%) | 47 (32.4%) | 36 (47.4%) |
| Previous TB | 37 (37.0%) | 54 (37.2%) | 42 (55.3%) |
| HIV positive | 35 (35.0%) | 49 (34.0%) | 23 (30.3%) |

### Sensitivity and specificity of TB testing platforms applied to tongue swab samples

Both Xpert Ultra and MTB Ultima exhibited better sensitivity in Cohort 2 than in Cohort 1 (**Table 2)**, though the difference in cohort positivity for either platform was not statistically significant (p = 0.21 and p = 0.08, respectively, by 2-tailed z score). When the results of both cohorts were combined, Xpert Ultra was 75.4% sensitive and 100% specific relative to sputum MRS. MTB Ultima was 71.6% sensitive and 96.9% specific relative to sputum MRS **(Table 2)**.

**Table 2:** Sensitivity and specificity of swab testing by Xpert Ultra and MTB Ultima, based on all samples tested.

| Co-hort | Sensitivity |  |  |  | Specificity |  |  |  |
| --- | --- | --- | --- | --- | --- | --- | --- | --- |
|  | Xpert Ultra |  | MTB Ultima |  | Xpert Ultra |  | MTB Ultima |  |
|  | N | Percent (95% CI) | N | Percent (95% CI) | N | Percent (95% CI) | N | Percent (95% CI) |
| 1 | 71/100 | 71.0 (61.1, 79.6) | 58/89 | 65.2 (54.3, 75.0) | -* | -* | -* | -* |
| 2 | 113/144 | 78.5 (70.9, 84.9) | 103/136 | 75.7 (67.7, 82.7) | 72/72 | 100.0 (95.0, 100.0) | 62/64 | 96.9 (89.2, 99.6) |
| 1+2 | 184/244** | 75.4 (69.5, 80.7) | 161/225** | 71.6 (65.2, 77.4) | 72/72** | 100.0 (95.0, 100.0) | 62/64** | 96.9 (89.2, 99.6) |

\*Cohort 1 consisted entirely of TB-positive participants by sputum MRS \*\*Totals fell below the total enrollment of 320 participants due to invalid results, which differed between the two test platforms

Due to instrument error messages, results deemed not valid after positive control runs (see Methods), or a lack of available samples for retesting, some results (N = 9 Xpert Ultra and 34 MTB Ultima) were excluded from the overall results in Table 2. Therefore, the Xpert Ultra and MTB Ultima sample sets in Table 2 do not precisely align. **Table 3** shows a smaller sample set (N = 287) that consists entirely of samples from Cohort 1 and Cohort 2 participants whose swabs were tested by both methods. In this paired analysis set, Xpert Ultra appeared to have slightly higher sensitivity than MTB Ultima. However, the difference was not statistically significant (p = 0.34 by 2-tailed z score).

**Table 3:** Sensitivity and specificity of swab testing by Xpert Ultra and MTB Ultima, based on samples from participants whose swabs were tested by both methods.

| Co-hort | Sensitivity |  |  |  | Specificity |  |  |  |
| --- | --- | --- | --- | --- | --- | --- | --- | --- |
|  | Xpert Ultra |  | MTB Ultima |  | Xpert Ultra |  | MTB Ultima |  |
|  | N | Percent (95% CI) | N | Percent (95% CI) | N | Percent (95% CI) | N | Percent (95% CI) |
| 1 | 63/89 | 70.8 (60.2, 80.0) | 58/89 | 65.2 (54.3, 75.0) | -* | -* | -* | -* |
| 2 | 107/136 | 78.7 (70.8, 85.2) | 103/136 | 75.7 (67.6, 82.7) | 62/62 | 100.0 (94.2, 100.0) | 60/62 | 96.8 (88.8, 99.6) |
| 1+2 | 170/225 | 75.6 (69.4, 81.0) | 161/225 | 71.6 (65.2, 77.4) | 62/62 | 100.0 (94.2, 100.0) | 60/62 | 96.8 (88.8, 99.6) |

### Tongue swab sensitivity varied with sputum Xpert Ultra semi-quantitative results

Sputum Xpert semi-quantitative data was available for a subset of Cohort 2 participants with sputum MRS-confirmed TB (N = 93). By both Xpert Ultra and MTB Ultima, tongue swab positivity was more common among patients who had higher sputum semi-quantitative results (**Table 4**).

**Table 4:** Sensitivity relative to sputum semi-quantitative results. Sputum semi-quantitative data were available for N = 93 participants with confirmed TB in Cohort 2.

| Sputum semi-quantitative results | Sensitivity, Xpert Ultra |  | Sensitivity, MTB Ultima |  |
| --- | --- | --- | --- | --- |
|  | N | Percent (95% CI) | N | Percent (95% CI) |
| High | 43/43 | 100 (91.8, 100) | 38/40 | 95 (83.1, 99.4) |
| Medium | 16/16 | 100 (79.4, 100) | 13/15 | 86.7 (59.5, 98.3) |
| Low | 16/23 | 69.6 (47.1, 86.8) | 13/21 | 61.9 (38.4, 81.9) |
| Very low | 3/8 | 37.5 (8.5, 75.5) | 4/7 | 57.1 (18.4, 90.1) |
| Trace | 0/3 | 0 (0, 70.8) | 0/3 | 0 (0, 70.8) |
| **Total** | **78/93** | **83.9 (74.8, 90.9)** | **68/86** | **79.1 (69, 87.1)** |

### Increased MTB Ultima sensitivity upon re-testing of swab sample lysates

Our swab sample processing protocol for MTB Ultima generated ~500 µL of crude lysate, of which only 6 µL was tested with the MTB Ultima chip. The remaining ~494 µL was returned to the -80 °C freezer. We asked whether we might see improved overall sensitivity if the frozen material was thawed and re-tested by running another 6 µL aliquot on MTB Ultima chips.

As shown in Table 2, the first-pass MTB Ultima analysis of 225 swab samples from participants with confirmed TB identified 161 swabs that were true-positive relative to sputum MRS, and 60 that were false-negative. Fifty-seven of the false-negative lysates were thawed and re-tested in fresh MTB Ultima chips. One of these samples yielded an invalid result. Of the 56 samples with valid results, 39 remained false-negative, while 17 yielded true-positive results on the second try. Positive swab results on the second run included “Very low” (N = 8), “Low” (N = 6), “Medium” (N = 2), and “High” (N = 1) MTB Ultima semi-quantitative values from swabs. When the 17 samples that became true-positive upon re-testing are added to the 161 samples that were true positive in the first pass, the adjusted sensitivity of MTB Ultima relative to sputum MRS increases to 178/225, or 79.1% (95% CI 73.2%, 84.2%).

In addition to false-negative samples, we also thawed and re-tested 28 true-negative samples (swabs from sputum MRS-negative participants that were negative by MTB Ultima on the first pass). All 28 of these samples (100%) yielded negative results upon re-testing.

## DISCUSSION

This study expanded on previous studies in three significant ways. It is among the first studies to evaluate dry collection/storage of tongue swab samples from TB patients. Dry swabs are likely to be easier to handle than buffer-filled tubes in the context of high-throughput community settings, which are often envisioned for non-sputum sampling (2-4, 18). Second, the current study used a streamlined protocol for swab sample processing for Xpert Ultra (about 15 minutes from sample tube to start of test, compared to 45 to 60 minutes for the methods described previously (6)). For occupational safety, an initial heat inactivation step (10 min at 100 °C) was conducted on the closed swab tube before it was opened for the first time. Third, we assessed a second commercial TB diagnostic NAAT test, MTB Ultima, that was applied to replicate tongue swabs collected from the same participants.

For Xpert Ultra testing of swabs, we found that the use of dry-stored swabs with a streamlined protocol resulted in a low error rate of 1%, with no loss of accuracy relative to previous methods that used wet-stored swabs and a lengthier protocol. The overall sensitivity of 75% observed here is similar to the previously reported sensitivity of 72% relative to sputum MRS and 78% relative to sputum Xpert Ultra (6). Specificity relative to sputum MRS was 100% in both studies. These similarities are notable given that the two studies were conducted in different settings and populations (Worcester, South Africa and Kampala, Uganda, respectively). While higher sensitivity relative to sputum MRS would be desirable and appears feasible based on results from studies that used manual methods (8, 14), the consistency observed between these two studies shows promise for the biological reproducibility of tongue swab sampling.

MTB Ultima testing of swabs yielded an error rate of 1.7% and an initial sensitivity value that was similar to Xpert Ultra testing. Within the subset of participants (Table 3) for whom side-by-side comparison of replicate swabs was possible, sensitivities relative to sputum MRS were 75.6% and 71.6% for Xpert Ultra and MTB Ultima, respectively. Specificity of both methods was high at >95%.

Both molecular tests exhibited higher sensitivities in Cohort 2 than in Cohort 1, though the differences were not significant. The two cohorts had different, but overlapping, time periods. Most Cohort 1 participants were enrolled in 2021, while most Cohort 2 participants with confirmed TB were enrolled in 2022 and 2023. The evolving COVID-19 pandemic may have affected the timing of care-seeking behaviors.

In our previous evaluation of Xpert Ultra analysis of tongue swabs (6), there appeared to be a relationship between tongue swab positivity and sputum swab Xpert Ultra semi-quantitative results. Positivity ranged from 100% (21/21) in patients with “high” sputum semi-quantitative results, down to 0% (0/8) in patients with “very low” or “trace” sputum results (6). Similarly, when stratified by sputum Xpert Ultra semi-quantitative results, both platforms used here were very sensitive in participants with higher semi-quantitative results, but less so in those with lower semi-quantitative results (**Table 4)**. These findings suggest that tongue swab testing for pauci-bacillary TB, such as would be expected in PLWHIV, children and mild subclinical disease, would need further optimization of sample collection and assay methods to improve performance relative to active TB with high bacillary burden.

Based on the results of our analysis of re-frozen and thawed lysates, there may be room for improvement in our MTB Ultima sample preparation protocol. Of 56 samples that were false-negative by MTB Ultima, 17 became true-positive upon re-testing. The reasons for this are not known. Sample contamination during re-testing does not seem likely given the 100% specificity observed with 28 re-tested true-negative samples. One possibility is that the additional freeze-thaw cycle may have further disrupted MTB bacilli that were damaged, but not disintegrated, in our initial bead-beading protocol. Freeze-thaw cycles are reported to enhance disruption of other bacteria (22). An alternative explanation is stochastic. Because our bead-beating protocol does not fully solubilize the material in swab samples, lysates have particulate cellular debris that may associate with MTB DNA in ways that are not uniform throughout the 500 µL suspensions. If so, then 6 µL aliquots applied to the Truenat chip may not always have representative amounts of target MTB DNA. Consistent with this possibility, of the 17 false-negative samples that became positive upon re-testing, 3 had MTB Ultima semi-quantitative signals that were quite robust on the second pass (semi-quantitative “Medium” or “High”). Thus, the effects of re-testing were non-uniform and greater than incremental in a few cases, consistent with a stochastic model. In view of these possibilities, improvements to the Truenat method could include enhanced mechanical disruptions (14, 23) and/or concentration of MTB DNA by methods such as magnetic particle hybridization capture (24, 25).

Strengths of our study included the side-by-side evaluation of two different molecular testing platforms. Moreover, our participant population included 245 participants with sputum MRS-confirmed TB, a much larger N than our previous study of Xpert Ultra applied to tongue swabs (6). There were several limitations. Because swabs evaluated in this study were collected in South Africa and tested in the USA, all of them underwent an initial freeze-thaw cycle for storage and shipment, a process that could have affected sensitivity in either direction. Sample preparation protocols for both tests required equipment (heat block, vortexer, and vortex adapter) which, although inexpensive, might not be available in all settings. A more powerful Biospec bead-beater was reported by Steadman et al (14) to yield excellent results when paired with a manual MTB qPCR applied to swabs, however the Biospec is expensive and not common outside of research laboratories. Finally, both commercial test platforms evaluated in this study were engineered to test sputum, not swabs, and there may be limitations to how well they can ever perform with swab samples.

Despite these limitations, our observations expand the evidence supporting tongue swabs as alternatives to sputum collection in settings where sputum collection is not possible. Tongue swabs are easier and faster to collect than any TB sample type that we are aware of, and their use could help increase access to TB testing and decrease occupational risks to healthcare workers. Further development of testing methods designed explicitly for TB tongue swabs, such as the high-volume qPCR approach described by Steadman et al (14), may deliver further improvements in accuracy relative to sputum testing. As we learned when anterior nasal sampling was introduced for COVID-19 testing (2, 3, 26), sample types that are non-invasive and easy to self-collect can be invaluable tools in the fight against transmissible diseases.

## Data Availability

All data produced in the present study are available upon reasonable request to the authors

## ACKNOWLEDGEMENTS

This work was supported by the Bill and Melinda Gates Foundation (#INV-004527, OPP 1213054), by NIH grant R01AI139254, and by the Australian Government. KAL was supported in part by a Mary Gates Undergraduate Research Fellowship. RC was supported in part by a University of Washington Department of Environmental and Occupational Health Sciences Top Scholar Award. We are grateful to Copan Italia for providing swabs. We are grateful to B.R. Sivakumar, Akron D’Souza, Winnie Gonsalves of Molbio, and Gayatri Chilambi of Rutgers University, for facilitating access to Molbio tests and instruments.

